# Validation of a clinical and genetic model for predicting severe COVID-19

**DOI:** 10.1101/2022.01.14.22269270

**Authors:** Gillian S. Dite, Nicholas M. Murphy, Erika Spaeth, Richard Allman, Lifelines Corona Research Initiative

**Affiliations:** Genetic Technologies Limited, Fitzroy, Victoria, Australia; Phenogen Sciences Inc, Charlotte, North Carolina, United States of America

**Keywords:** severe COVID-19, risk prediction, validation, risk factors, single-nucleotide polymorphism

## Abstract

Using nested case–control data from the Lifelines COVID-19 cohort, we undertook a validation study of a clinical and genetic model to predict the risk of severe COVID-19 in people with confirmed COVID-19 and in people with confirmed or self-reported COVID-19. The model performed well in terms of discrimination of cases and controls for all ages (area under the receiver operating characteristic curve [AUC] = 0.680 for confirmed COVID-19 and AUC = 0.689 for confirmed and self-reported COVID-19) and in the age group in which the model was developed (50 years and older; AUC = 0.658 for confirmed COVID-19 and AUC= 0.651 for confirmed and self-reported COVID-19). There was no evidence of over- or under-dispersion of risk scores but there was evidence of overall over-estimation of risk in all analyses (all *P* < 0.0001). In the light of large numbers of people worldwide remaining unvaccinated and continuing uncertainty regarding vaccine efficacy over time and against variants of concern, identification of people at high risk of severe COVID-19 may encourage the uptake of vaccinations (including boosters) and the use of non-pharmaceutical inventions.

## Text

Severe coronavirus disease 2019 (COVID-19) disproportionately affects older adults, but can occur in people of all ages, especially those with comorbidities [1]. An abundance of research has identified clinical and genetic risk factors that are associated with developing severe disease if infected with severe acute respiratory syndrome-coronavirus-2 (SARS-CoV-2) [1, 2]. In clinical practice, information on these risk factors can be useful when combined in a risk prediction model that provides a single estimate of absolute risk that enables health care providers to effectively communicate with their patients about risk.

We previously described a clinical and genetic model for predicting severe COVID-19 that was developed and validated using data from the UK Biobank [3]. We now report the results of a validation of the model using a case–control analysis of an external dataset from the Netherlands [4]. While the risk prediction model was developed in people aged 50 years and older, here we assess its performance in people aged 24 years and older.

We used data from participants in the Lifelines COVID-19 cohort [4] who were recruited from the Lifelines and Lifelines NEXT cohorts [5, 6]. Lifelines is a multi-disciplinary prospective population-based cohort study examining, in a unique three-generation design, the health and health-related behaviours of 167,729 people living in the north of the Netherlands. Lifelines employs a broad range of investigative procedures in assessing the biomedical, socio-demographic, behavioural, physical and psychological factors that contribute to the health and disease of the general population, with a special focus on multi-morbidity and complex genetics.

During 2020, questionnaire links were emailed to Lifelines and Lifelines NEXT participants, weekly from the end of March to mid-May and then every two weeks until July, after which the questionnaires were sent monthly through to April 2021 [4]. Lifelines COVID-19 cohort participants were aged 24 years or over and had completed at least one of the regular online COVID-19 questionnaires via an emailed link during the first eight weeks of data collection [4].

The questionnaire response dates corresponded to the period from around one month after the beginning of the first wave of the COVID-19 pandemic in the Netherlands through to the peak of the fourth wave in May 2021. During this time, the original SARS-CoV-2 virus accounted for over 95% of infections in the Netherlands until early January 2021, after which the alpha variant became more prevalent and had accounted for over 95% of infections by the end of March 2021 [7]. The presence of the delta variant was negligible during the period of data collection for this study.

COVID-19 vaccinations became available in the Netherlands in mid-January 2021 and were initially offered to high-risk groups and then progressively to other groups (such as care workers) and younger age groups until all adults became eligible in mid-June 2021 [8]. From questionnaire 18 (March 2021) onwards, participants were asked about their vaccination status and we excluded questionnaires where (and after which) a participant reported having had one or two doses of a vaccine.

At the beginning of data collection, when testing for SARS-CoV-2 infection was not widely available in the Netherlands, Lifelines COVID-19 questionnaires 1–4 asked participants whether a doctor had told them they had COVID-19 [4]. From questionnaire 5 (early May 2020) onwards, the questionnaires also asked about positive test results. From these questions we identified a group of participants with confirmed COVID-19. In addition, the questionnaires asked participants to self-report having had COVID-19. We used this question with the previous questions to identify a broader group of participants who had either confirmed or self-reported COVID-19.

Given the limited availability of testing early in the data collection period, the confirmed COVID-19 group is likely to miss some participants who had COVID-19. Conversely, the broader group including participants with self-reported COVID-19 is likely to have some false positives. The true number of participants who had COVID-19 will be somewhere between the two. Therefore, we conducted two sets of analyses: (i) using participants with confirmed COVID-19 and (ii) using participants with confirmed and self-reported COVID-19.

As we did previously, we used hospitalization as a proxy for severe COVID-19 [3]. The Lifelines COVID-19 questionnaires specifically asked participants whether they had been hospitalized for COVID-19. The questionnaires also asked about being given supplemental oxygen, admission to an intensive care unit and being placed on a ventilator, but there were too few positive responses to these questions to allow separate analysis.

The risk factors included in the calculation of the risk of severe COVID-19 are age; sex; body mass index; a history of cerebrovascular disease, diabetes, haematological cancer, non-haematological cancer, hypertension, kidney disease or respiratory disease (excluding asthma); and the genotypes of seven single nucleotide polymorphisms (SNPs) – rs112641600, rs10755709, rs118072448, rs7027911, rs71481792, rs112317747 and rs2034831 [3]. The log odds of the risk of severe COVID-19 is the sum of the intercept and the product of the value and beta coefficient for each of the risk factors listed in Supplementary Table S1. The probability of severe COVID-19 is then the inverse logit of the log odds (*x*), that is, 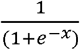.

We used the age reported at the completion of the participant’s first Lifelines COVID-19 questionnaire. The questionnaires asked about a history of cancer, cerebrovascular disease, diabetes, hypertension, kidney disease and respiratory disease on three occasions. If any of the participants’ responses to the risk factor questions were missing for all answered questionnaires, we used responses from their Lifelines baseline questionnaire.

We were not able to identify the type of cancer reported by Lifelines COVID-19 participants so we used the risk associated with having a non-haematological cancer for all reported cancers. In the Lifelines questionnaires, the respiratory disease question included asthma, whereas this is excluded in the model calculations. Because we were not able to distinguish respiratory disease solely due to asthma, we included all reports of respiratory disease in the model calculations. Gender, ethnicity, weight and height were taken from the Lifelines baseline questionnaire. If two weight or height measurements were available, we used the most recent weight measurement and the mean of the height measurements. Two of the SNPs in the risk model were not available on the Illumina CytoSNP-12v2 array used by Lifelines [6]. Instead, we used highly correlated proxy SNPs (rs10905502 was the proxy for rs71481792 [r^2^ = 0.75, D’ = 1.0] and rs78654835 was the proxy for rs112317747 [r^2^ = 1.0, D’ = 1.0]).

To extend the model to people aged less than 50 years, we estimated the risk associated with younger age groups using data from the Centers for Disease Control and Prevention [9] such that, compared with the 50–69 years baseline age group, people aged 18– 29 years were at 0.27 times the risk, people aged 30–39 years were at 0.43 times the risk, and people aged 40–49 years were at 0.67 times the risk.

In each analysis – (i) using participants with confirmed COVID-19 and (ii) using participants with both confirmed and self-reported COVID-19 – the cases were those who reported having been hospitalized for COVID-19 and the controls were the remainder of the group. We also did analyses restricting the dataset to those aged 50 years or older (the ages in which the model was developed).

As we did previously [3], we assessed the association between quintile of risk score and severe COVID-19 using logistic regression. We used the area under the receiver operating characteristic curve (AUC) to assess discrimination. We used logistic regression of the log odds of the risk score to assess calibration in terms of the overall estimation of risk (the intercept) and the dispersion of risk (the slope), and we drew calibration plots of deciles of expected and observed cases of severe COVID-19. We used Stata MP version 13.1 (StataCorp LP, College Station, Texas, USA) for all analyses and all statistical tests were two sided.

The Lifelines protocol has been approved by the Medical Ethical Committee of the University Medical Center Groningen, The Netherlands, under Approval Number 2007/152. All participants provided written informed consent to Lifelines before data collection began. This research was conducted using Lifelines data under Project Number OV20-00101.

The data used in this study was made available to us by Lifelines and is not publicly available. Researchers can apply to use the Lifelines data used in this study, and more information about how to request Lifelines data and the conditions of use can be found on their website (https://www.lifelines.nl/researcher/how-to-apply). Stata MP Version 13.1 code for the analysis is available for non-commercial purposes from the corresponding author on request.

Of the 26 845 Lifelines COVID-19 cohort participants who had genotyping data available and had completed at least one questionnaire, 3214 (12.0%) completed one questionnaire, 5742 (21.4%) completed 2–5 questionnaires, 4194 (15.6%) completed 6–10 questionnaires, 3568 (13.3%) completed 11–15 questionnaires, 6106 (22.7%) completed 16– 20 questionnaires and 4021 (15.0%) completed 21–23 questionnaires. We excluded 15 933 questionnaires from 15 040 participants where (and after which) the participant reported being vaccinated.

In the final dataset, 55 participants were hospitalized for their COVID-19 infection and were considered cases in this study. We used two control groups: the first comprised the 1355 participants who had confirmed COVID-19; the second comprised both the first control group and the 2518 participants who self-reported having had COVID-19 (i.e. 3925 participants). In the cases, there were 28 (50.9%) women and 27 (49.1%) men; their mean age was 57.6 years (standard deviation [SD] = 10.3) and the mean number of completed questionnaires was 17.0 (SD = 6.3). In the confirmed COVID-19 control group, there were 905 (66.8%) women and 450 (33.2%) men; their mean age was 53.0 years (SD = 11.6) and the mean number of completed questionnaires was 14.1 (SD = 6.8). In the confirmed and self-reported COVID-19 control group, there were 2414 (62.3%) women and 1459 (37.7%) men; their mean age was 51.5 years (SD = 11.8) and the mean number of completed questionnaires was 12.2 (SD = 7.2).

In the cases, the mean probability of severe COVID-19 was 0.225 (SD = 0.019); in the confirmed COVID-19 controls, the mean was 0.165 (SD = 0.002); and in the confirmed and self-reported COVID-19 controls, the mean was 0.165 (SD = 0.001). The risk distribution for the cases, both control groups and the whole Lifelines COVID-19 cohort are show in in Supplementary Figure S1.

The top half of Table 1 shows the results of the analyses of the confirmed COVID-19 group and the confirmed and self-reported COVID-19 group for all ages. Overall, the results were similar for the two groups. The odds ratios (OR) per quintile of risk (1.63 and 1.58, respectively) were a little lower than the OR of 1.77 seen in the validation group in the risk prediction model development paper [3]. Similarly, the AUCs (0.680 and 0.679, respectively) were a little lower than the AUC of 0.732 seen in the model development paper. In terms of calibration, there was no evidence of under- or over-dispersion in either group (β = 0.92 and 0.90, respectively), as in the original paper (β = 0.90). In both groups, the model overestimated risk (α = −1.78 and −2.86, respectively), whereas the validation group in the model development paper did not (α = −0.08).

**Table 1.** Validation analysis of model to predict risk of severe COVID-19 for participants of all ages and for participants aged 50 years and older

| All ages | Estimate | 95% CI | P value |
| --- | --- | --- | --- |
| **Association: OR per quintile of risk** |  |  |  |
| Confirmed COVID-19 | 1.63 | 1.31, 2.04 | <0.0001 |
| Confirmed and self-reported COVID-19 | 1.58 | 1.27, 1.95 | <0.0001 |
| **Discrimination: AUC** |  |  |  |
| Confirmed COVID-19 | 0.680 | 0.608, 0.752 |  |
| Confirmed and self-reported COVID-19 | 0.679 | 0.607, 0.751 |  |
| **Calibration slope: $\beta$** |  |  |  |
| Confirmed COVID-19 | 0.92 | 0.54, 1.30 | 0.7* |
| Confirmed and self-reported COVID-19 | 0.90 | 0.55, 1.25 | 0.6* |
| **Calibration intercept: $\alpha$** |  |  |  |
| Confirmed COVID-19 | -1.78 | -2.38, -1.19 | <0.0001 |
| Confirmed and self-reported COVID-19 | -2.86 | -3.41, -2.30 | <0.0001 |
| Aged 50 years and older | Estimate | 95% CI | P value |
| **Association: OR per quintile of risk** |  |  |  |
| Confirmed COVID-19 | 1.54 | 1.21, 1.96 | <0.0001 |
| Confirmed and self-reported COVID-19 | 1.47 | 1.16, 1.86 | 0.001 |
| **Discrimination: AUC** |  |  |  |
| Confirmed COVID-19 | 0.658 | 0.579, 0.737 |  |
| Confirmed and self-reported COVID-19 | 0.651 | 0.573, 0.730 |  |
| **Calibration slope: $\beta$** |  |  |  |
| Confirmed COVID-19 | 0.75 | 0.34, 1.16 | 0.2* |
| Confirmed and self-reported COVID-19 | 0.69 | 0.31, 1.08 | 0.1* |
| **Calibration intercept: $\alpha$** |  |  |  |
| Confirmed COVID-19 | -1.85 | -2.48, -1.21 | <0.0001 |
| Confirmed and self-reported COVID-19 | -2.87 | -3.46, -2.28 | <0.0001 |

Note: AUC, area under the receiver operating characteristic curve; CI, confidence interval; OR, odds ratio. \* P value for the null hypothesis that the calibration slope = 1.

The bottom half of Table 1 shows the analyses limited to participants aged 50 years and older. Compared with the analysis of all ages, there was a reduction in the ORs per quintile of risk for both the confirmed COVID-19 group and the confirmed and self-reported COVID-19 group (1.54 and 1.47, respectively), and a reduction in the AUCs (0.658 and 0.651, respectively). The calibration slopes suggested over-dispersion of risk but were not statistically significant. The overestimations of risk (−1.84 and −2.87, respectively) were similar to those seen in the analysis of all ages.

The true number of people with COVID-19 is unknown but is likely to be somewhere between the number who test positive for SARS-CoV-2 infection and the number who self-report having had COVID-19. In this study we have addressed this uncertainty by conducting two sets of analyses: the first in individuals with confirmed COVID-19, and the second with individuals with confirmed and self-reported COVID-19. In terms of discrimination, the AUC of the risk prediction model was almost identical in the two analyses and only slightly lower than the AUC in validation group in the model development paper [3]. This and the similarity in the association per quintile of risk (Table 1) provide confidence in the model’s application across adult populations. Risk of COVID-19 severity was overestimated in this study, but in a clinical setting, overestimation of risk is preferred to an underestimation given that the risk-reduction options of vaccination, masking and social distancing are benign in nature. Our results were similar for the full dataset and when limiting analyses to people aged 50 years and over.

It is possible that some severe cases of COVID-19 have not been ascertained in this dataset. Death registry linkage identified 77 deaths in the broader Lifelines COVID-19 cohort in people who did not have confirmed or self-reported COVID-19. While these deaths may have been unrelated to COVID-19, some will represent people who became infected and were too unwell to complete a Lifelines COVID-19 questionnaire before they died. This limitation may have attenuated some of the results seen in this study.

As the pandemic continues to evolve, there are two major issues that can affect the utility of our risk model. First, we have to address the impact of viral variants on the performance of the risk model. The model development paper [3] and the present study used datasets in which the original and alpha SARS-CoV-2 variants were predominant. We have not been able to assess our model in datasets with known delta or omicron SARS-CoV-2 variants. We hypothesize that the clinical and genetic risk factors have broad effects in terms of risk of severe disease because the delta and omicron SARS-CoV-2 variants appear to affect transmissibility rather than severity [10].

Second, our model does not incorporate the protection offered by vaccination. Thus in vaccinated adults, the model will overestimate their risk of developing severe disease. However, we know that vaccine immunity wanes over about six months through a steady reduction in antibody levels leading to greater number of breakthrough infections among the vaccinated [11]. The wide range of immunity across individuals makes it hard to predict the impact of waning vaccination in terms of risk. Thus, we believe that the model can be used to provide a baseline risk of developing severe disease, even in the context of vaccinated adults.

Herein, we have validated our model to predict risk of severe COVID-19 if infected with SARS-CoV-2 in a dataset unrelated to the one in which the model was originally developed and validated. Despite new SARS-CoV-2 variants of concern, the model may complement current public health efforts in vaccine (and booster) uptake and may enable healthcare providers to have more informed discussions with patients about their risk-mitigation options and early treatment awareness, if ever infected.

## Data Availability

The data used in this study was made available to us by Lifelines and is not publicly available. Researchers can apply to use the Lifelines data used in this study, and more information about how to request Lifelines data and the conditions of use can be found on their website (https://www.lifelines.nl/researcher/how-to-apply).

## Acknowledgements

The authors wish to acknowledge the services of the Lifelines Cohort Study (especially the data management team), the contributing research centres delivering data to Lifelines and all the study participants.

## Supplementary information

**Supplementary Table S1.** Beta coefficients for calculation of risk of severe COVID-19

| Variable | Value | Beta coefficient |
| --- | --- | --- |
| Intercept |  | -1.37 |
| Age 18–29 years | 0 = no, 1 = yes | -1.31 |
| Age 30–39 years | 0 = no, 1 = yes | -0.83 |
| Age 40–49 years | 0 = no, 1 = yes | -0.40 |
| Age 50–69 years | 0 = no, 1 = yes | 0.00 |
| Age 70–74 years | 0 = no, 1 = yes | 0.57 |
| Age 75–79 years | 0 = no, 1 = yes | 0.82 |
| Age 80+ years | 0 = no, 1 = yes | 1.01 |
| Male | 0 = no, 1 = yes | 0.24 |
| Inverse of BMI | 10/BMI | -1.60 |
| Cancer, haematological | 0 = no, 1 = yes | 1.00 |
| Cancer, non-haematological | 0 = no, 1 = yes | 0.26 |
| Cerebrovascular disease | 0 = no, 1 = yes | 0.40 |
| Diabetes | 0 = no, 1 = yes | 0.43 |
| Hypertension | 0 = no, 1 = yes | 0.29 |
| Kidney disease | 0 = no, 1 = yes | 0.69 |
| Respiratory disease (excluding asthma) | 0 = no, 1 = yes | 1.17 |
| rs112317747 | 0 = T/T, 1 = C/T, 2 = C/C | 0.27 |
| rs2034831 | 0 = A/A, 1 = C/A, 2 = C/C | 0.24 |
| rs112641600 | 0 = C/C, 1 = T/C, 2 = T/T | -0.24 |
| rs10755709 | 0 = A/A, 1 = G/A, 2 = G/G | 0.12 |
| rs118072448 | 0 = T/T, 1 = C/T, 2 = C/C | -0.20 |
| rs7027911 | 0 = G/G, 1 = A/G, 2 = A/A | 0.10 |
| rs71481792 | 0 = A/A, 1 = T/A, 2 = T/T | -0.11 |

Note: Body mass index (BMI) is calculated as kg/m<sup>2</sup>; the inverse of BMI is calculated as 10 divided by BMI. In the current analysis, we used rs10905502 as a proxy for rs71481792 and rs78654835 as a proxy for rs112317747.

**Supplementary Figure S1.**
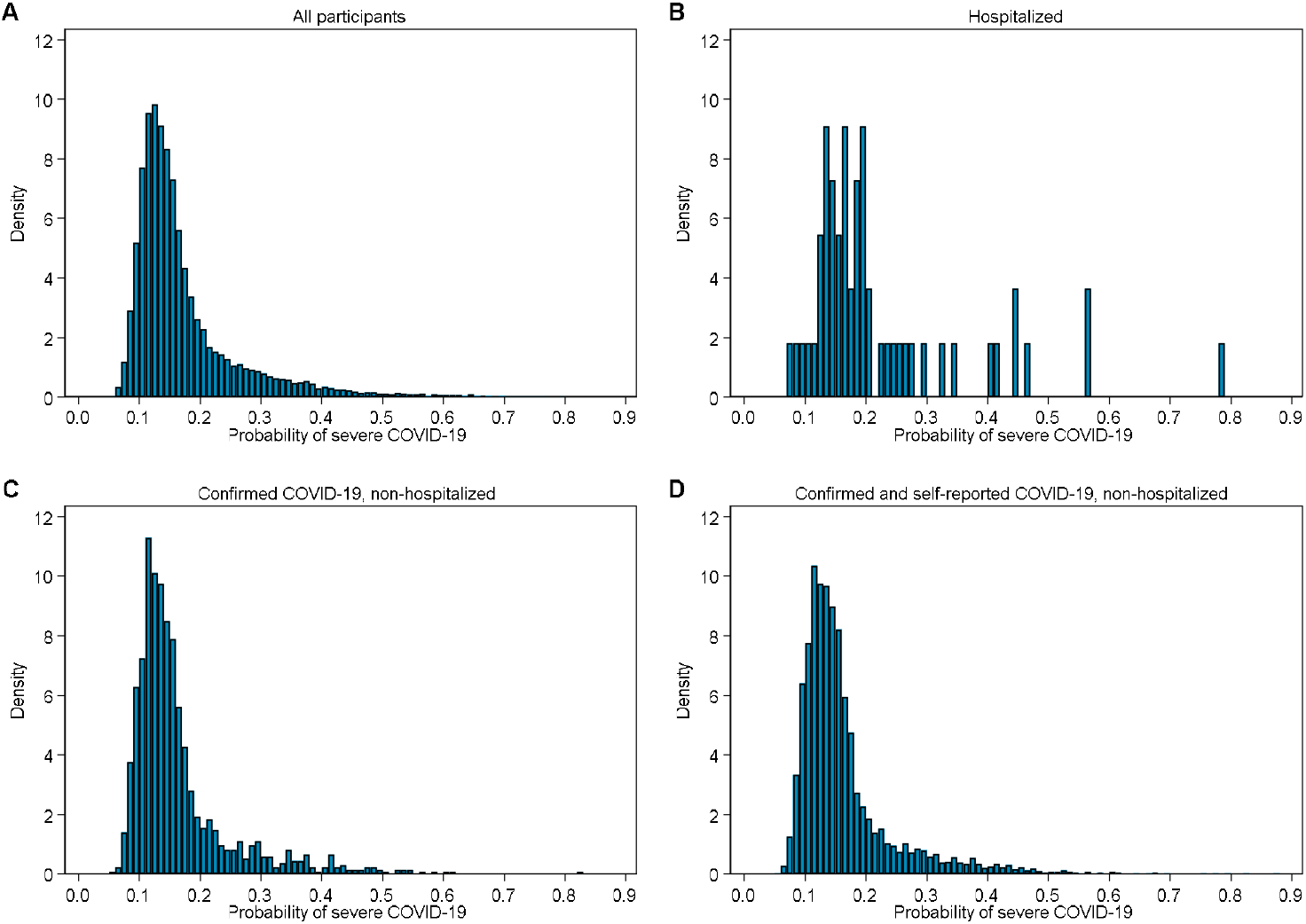
Distribution of probability of severe COVID-19 in (A) all Lifelines COVID-19 cohort participants, (B) hospitalized (cases), (C) non-hospitalized confirmed COVID-19 (controls) and (D) non-hospitalized confirmed and self-reported COVID-19 (controls).

**Supplementary Figure S2.**
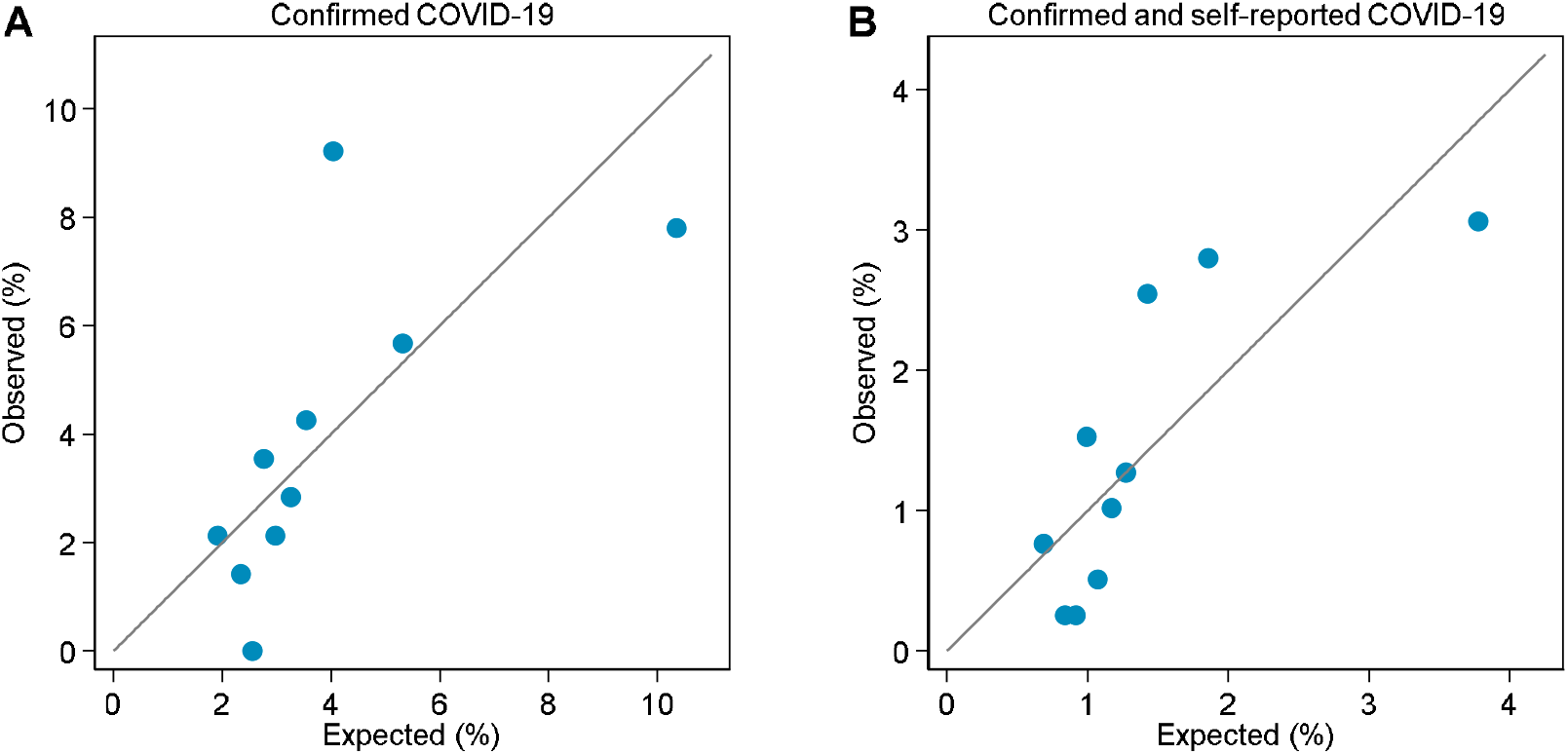
Calibration plots for (A) the confirmed COVID-19 group and (B) the confirmed and self-reported COVID-19 group.

